# Characteristics associated with frequent sexually transmitted infection (STI) testing in a community-based sample of gay, bisexual, and other men who have sex with men (GBMSM), United Kingdom, 2024

**DOI:** 10.1101/2025.10.07.25337534

**Authors:** Lucy Findlater, George Baldry, Ana K. Harb, Dolores Mullen, Dawn Phillips, Erna Buitendam, Catherine M. Lowndes, David Reid, Catherine H. Mercer, John Saunders, Dana Ogaz, Hamish Mohammed

**Author notes:** Corresponding author: (LF).

## Abstract

In the UK, gay, bisexual, and other men who have sex with men (GBMSM) at risk of sexually transmitted infections (STIs) are recommended quarterly testing, but it is not known how many are following this recommendation. We described the prevalence and correlates of frequent STI testing amongst GBMSM. We analysed data on participants of the community-based Reducing Inequalities in Sexual Health (RiiSH) online survey of GBMSM, recruited in November-December 2024 via social media and dating apps. Participants were UK residents aged ≥16 years who reported sex with a man in the previous year. We described the frequency of STI testing amongst individuals recommended quarterly testing, using proxies for national guidelines for quarterly testing eligibility (over the past three months: new male sexual partner, condomless anal sex, ≥10 male partners, or chemsex). We explored factors associated with frequent testing (≥4 tests/past year) using univariate and multivariable logistic regression, adjusting for country of birth and residence, ethnicity, employment, and education. Among 2758 participants (median age 45 years, 88% white), we estimated that 2366 (86%) would be recommended quarterly STI testing. Over the past year, 562 individuals (24%) met or exceeded this recommendation (testing ≥4 times), 1107 (47%) had 1-3 tests, and 673 (28%) no tests. Factors associated with frequent testing were: reporting using PrEP in the past year (adjusted odds ratio 7.66 (95% confidence intervals 5.77-10.30)), STI diagnosis in the past three months (1.96 (1.45-2.64)), and younger age (1.50 (1.04-2.16), aged 16-29 years vs ≥45 years). Straight/bisexual orientation was associated with less frequent testing than gay/homosexual (0.71 (0.52-0.96)). Overall, data from a large UK community survey suggest that only 1 in 4 GBMSM who are recommended frequent STI testing meet this recommendation, and testing levels vary by PrEP use, STI history, age, and sexual orientation. Interventions are needed to address these inequalities.

## Introduction

Gay, bisexual, and other men who have sex with men (GBMSM) have disproportionately high rates of diagnosed sexually transmitted infections (STIs) [1, 2]. Rates of STIs are rising worldwide, with increases in the bacterial STIs syphilis, gonorrhoea, and chlamydia observed amongst GBMSM in high income countries [3].

Public health guidelines in most high-income countries recommend that GBMSM test at least annually for STIs and HIV, and that those engaging in behaviours associated with STI or HIV transmission should test more frequently [4–6]. In the United Kingdom (UK), it is recommended that all GBMSM test for STIs and HIV annually, and that GBMSM at increased risk of these infections test quarterly [7, 8]. Additionally, pre-exposure prophylaxis for HIV (PrEP), which has been routinely provided at sexual health services in all UK nations since autumn 2020, is usually available on a three month prescription from sexual health services, and quarterly (three-monthly) HIV testing is recommended as part of PrEP care continuum [9]. Together, these factors are thought to facilitate quarterly STI testing in GBMSM. In the UK, STI testing for GBMSM includes asymptomatic ‘triple site’ (pharyngeal, rectal, and urethral) screening for chlamydia and gonorrhoea [10, 11]. STI testing in the UK is provided through open-access sexual health services which are free at the point of delivery and include both in-person clinics and online postal self-sampling services for STIs.

Frequent STI testing is recommended to detect and treat asymptomatic infections which could act as a reservoir for onwards transmission [10, 12–14]. Studies have indicated that frequent STI screening amongst GBMSM using PrEP may be associated with reductions in gonorrhoea or chlamydia incidence [15]. However, frequent screening and treatment of asymptomatic chlamydia and gonorrhoea in GBMSM could also lead to increased emergence of antimicrobial resistance without benefit to the individual [16–18]. Asymptomatic infection with chlamydia or gonorrhoea in men can self-resolve and may not be associated with negative health consequences [19–21]. Additionally, actual STI testing frequency amongst GBMSM may be lower than recommended [22, 23].

To inform testing guidelines and research into the barriers to and facilitators of frequent STI testing, it is important to understand the actual frequency and correlates of STI testing amongst GBMSM. This paper seeks to do so using data on demographics, clinical outcomes, STI testing, and service utilisation reported by a large community sample of GBMSM in the 2024 round of the ‘Reducing Inequalities in Sexual Health’ (RiiSH) survey, which was administered from November to December 2024.

## Methods

### Study population

UK residents who were aged at least 16 years and reported sex in the past 12 months with a man (cisgender or transgender) or non-binary person assigned male at birth, participating in the 2024 round of the RiiSH survey.

### Study design

Secondary analysis of community-based, cross-sectional, online survey data.

### Data collection

Data collection methods for the RiiSH surveys have been described previously [24–30]. Participants were those who responded to an advert on social networking sites (Facebook, Instagram) or on geospatial dating applications (Grindr, Scruff, Jack’d, and Recon) between 19 November 2024 and 9 December 2024. Eligible individuals were UK residents, aged 16 years or older, men (cisgender or transgender), transgender women, or gender-diverse and assigned male at birth, who reported sex in the past 12 months with a man (cisgender or transgender) or non-binary person assigned male at birth.

The study used an anonymous, self-completed, online questionnaire administered via Snap Surveys platform (www.snapsurveys.com), taking approximately 15 minutes to complete. Online consent was obtained from participants and no financial incentive was offered. Young people aged 16–17 years were eligible to participate in this survey as this group represent a key population at risk of STI acquisition in the UK and are important to include in this research. Parental consent of participants aged 18 or younger was not sought. Ethical guidelines produced by the British Psychological Society and General Medical Council suggest consent from parents should be sought for those under age 16 and those aged 16 and over may be presumed to be able to reach informed consent if the information on the study, and the way that data is collected, stored, and used is clear. This information was included for all potential participants at the beginning of the survey. Question topics included HIV/STI testing, vaccination, PrEP use, STI symptoms, use of sexual health services, sexual relationships and behaviour, and drug and alcohol use.

### Data analysis

Data analysis was conducted using R version 4.2.1. Data were accessed for the purpose of this study on 17 February 2025. Authors were not able to identify participants during or after data collection because participants did not provide information which could identify them in the survey.

We assessed whether a participant would be recommended quarterly testing using UK national guidelines for the sexual health care of GBMSM [7, 8]. These guidelines recommend that quarterly STI and HIV testing should be offered to people with any of the following risk factors for STIs and bloodborne viruses: multiple or anonymous sexual partners since last tested, condomless anal sex (CAS) with partners of unknown or serodiscordant HIV status over the past year, any unprotected sexual contact with a new partner since last tested, or drug use during sex (chemsex) over the past six months. We used the following variables to approximate these criteria: any new male sexual partner, CAS, at least 10 male sexual partners, or chemsex, in the past three months. The threshold of at least 10 partners reflects the long-tailed distribution of testing in the dataset (S1 Figure). We considered using a lower threshold of at least 5 partners and performed a sensitivity analysis using this threshold, but we did not include this as it led to almost no difference (<10 additional people) in the sample of participants we would consider eligible for quarterly testing.

Amongst participants who would be eligible for the quarterly testing recommendation, we described the self-reported frequency of STI testing, defined as number of tests taken in the past year. A variable as a proxy for quarterly STI screening (yes or no) was created, defined as testing at least four times in the past year.

We described where the participant had ever undergone STI testing (e.g., sexual health services, self-testing, hospital, GP, community testing services), and if they had ever received an HIV test, if this was within the last year, and where the last HIV test was received (e.g., sexual health services, self-testing, hospital, GP, community testing services). We also calculated the frequency of STI symptoms and diagnoses amongst this group in the past three months, as well as reasons for visiting sexual health services.

To examine the association between frequent STI testing and sociodemographic and behavioural characteristics, univariable and multivariable logistic regression models were used to produce unadjusted (uOR) and adjusted odds ratios (aOR). Only participants with complete data for each variable in the model were included. We used a statistical approach to decide which variables to include in the multivariable model (using a threshold of p value ≤ 0.2 in univariable analysis for inclusion in multivariable) as well as evidence from the literature to identify variables with evidence of association with STI testing frequency, for example, age, ethnicity, STI history, and sexual orientation [25, 27].

The group of participants who did not report testing quarterly included people who reported not having had an STI test in the past year or ever. This meant that our analysis could be identifying factors associated with having any testing at all or having ever engaged with sexual health services, as opposed to factors associated with frequent testing. To explore this, we conducted a sensitivity analysis comparing people who tested at least four times in the past year to those who tested between one and three times in the past year. We used the same approach as described in the previous paragraph to build a multivariable logistic regression model and assess which variables were associated with testing at least four times in the past year as opposed to testing one, two, or three times.

## Results

### Sample characteristics

Overall, 2758 participants were included in RiiSH 2024, with a median age of 45 years, the majority of white ethnicity (88%), and identifying as gay or homosexual (80%) (Table 1).

**Table 1:** Characteristics of RiiSH participants overall and of RiiSH participants who would be recommended quarterly STI testing, 2024. Characteristics of participants in the Reducing Inequalities in Sexual Health (RiiSH) survey 2024, stratified based on whether they would be recommended quarterly testing for sexually transmitted infections (STIs) according to proxy eligibility criteria (any new male sexual partner, condomless anal sex, at least 10 male sexual partners, or chemsex, in the past three months). PrEP: pre-exposure prophylaxis for HIV. PLWHIV: person living with HIV.

| Characteristic | Whole sample (N = 2758) | Recommended quarterly testing (N = 2366) | Not recommended quarterly testing (N = 392) |
| --- | --- | --- | --- |
| Age (years) | Median 45 (IQR 36 – 55) | Median 45 (IQR 36 – 55) | Median 45 (IQR 35 – 56) |
| Age group (years) |  |  |  |
| 16-29 | 337 (12%) | 275 (12%) | 62 (16%) |
| 30-44 | 1,010 (37%) | 878 (37%) | 132 (34%) |
| 45 and over | 1,411 (51%) | 1,213 (51%) | 198 (51%) |
| Gender |  |  |  |
| Cisgender male | 2,631 (95%) | 2,272 (96%) | 359 (92%) |
| Other gender | 127 (4.6%) | 94 (4.0%) | 33 (8.4%) |
| Sexual orientation |  |  |  |
| Gay/homosexual | 2,207 (80%) | 1,908 (81%) | 299 (76%) |
| Bisexual, straight/heterosexual, or other | 551 (20%) | 458 (19%) | 93 (24%) |
| Country |  |  |  |
| England | 2,404 (87%) | 2,064 (87%) | 340 (87%) |
| Scotland | 194 (7.0%) | 165 (7.0%) | 29 (7.4%) |
| Wales | 100 (3.6%) | 82 (3.5%) | 18 (4.6%) |
| Northern Ireland | 60 (2.2%) | 55 (2.3%) | 5 (1.3%) |
| Region within England |  |  |  |
| East Midlands | 168 (6.1%) | 134 (5.7%) | 34 (8.7%) |
| East of England | 206 (7.5%) | 173 (7.3%) | 33 (8.4%) |
| London | 748 (27%) | 663 (28%) | 85 (22%) |
| North East | 89 (3.2%) | 74 (3.1%) | 15 (3.8%) |
| North West | 316 (11%) | 276 (12%) | 40 (10%) |
| South East | 323 (12%) | 285 (12%) | 38 (9.7%) |
| South West | 194 (7.0%) | 165 (7.0%) | 29 (7.4%) |
| West Midlands | 162 (5.9%) | 125 (5.3%) | 37 (9.4%) |
| Yorkshire and Humber | 189 (6.9%) | 161 (6.8%) | 28 (7.1%) |
| Unknown | 9 (0.3%) | 8 (0.3%) | 1 (0.3%) |
| Ethnicity |  |  |  |
| White | 2,435 (88%) | 2,094 (89%) | 341 (87%) |
| Asian | 141 (5.1%) | 119 (5.0%) | 22 (5.6%) |
| Mixed or other | 125 (4.5%) | 107 (4.5%) | 18 (4.6%) |
| Black | 54 (2.0%) | 43 (1.8%) | 11 (2.8%) |
| Unknown | 3 (0.1%) | 3 (0.1%) | 0 (0%) |
| Education |  |  |  |
| Degree or higher | 1,628 (59%) | 1,443 (61%) | 185 (47%) |
| Higher education below degree | 310 (11%) | 271 (11%) | 39 (9.9%) |
| A-levels, Scottish Highers, equivalent | 292 (11%) | 238 (10%) | 54 (14%) |
| GCSEs, O-Levels, National 5, equivalent | 380 (14%) | 305 (13%) | 75 (19%) |
| No qualifications | 70 (2.5%) | 46 (1.9%) | 24 (6.1%) |
| Other | 78 (2.8%) | 63 (2.7%) | 15 (3.8%) |
| Employed | 2,139 (78%) | 1,863 (79%) | 276 (70%) |
| Used PrEP in last year |  |  |  |
| Yes | 1,248 (45%) | 1,172 (50%) | 76 (19%) |
| No | 1,234 (45%) | 957 (40%) | 277 (71%) |
| N/A | 276 (10%) | 237 (10%) | 39 (9.9%) |
| HIV status |  |  |  |
| Negative/unknown | 2,458 (89%) | 2,107 (89%) | 351 (90%) |
| PLWHIV (tested positive) | 300 (11%) | 259 (11%) | 41 (10%) |
| STI positive since August 2023 | 284 (10%) | 279 (12%) | 5 (1.3%) |
| Number of sexual partners since August 2024 |  |  |  |
| No sex/only virtual | 247 (9.0%) | 1 (<0.1%) | 246 (63%) |
| 1 | 309 (11%) | 216 (9.1%) | 93 (24%) |
| 2-4 | 844 (31%) | 799 (34%) | 45 (11%) |
| 5-9 | 607 (22%) | 600 (25%) | 7 (1.8%) |
| 10 or more | 751 (27%) | 750 (32%) | 1 (0.3%) |
| Any new partner since August 2024 |  |  |  |
| Had multiple partners and more than one partner was new | 1,784 (65%) | 1784 (75%) | 0 (0%) |
| Had multiple partners and one was new | 250 (9.1%) | 250 (11%) | 0 (0%) |
| Had only one partner who was also new | 112 (4.1%) | 112 (4.7%) | 0 (0%) |
| No new partners | 612 (22%) | 220 (9.3%) | 392 (100%) |

**Table 2:**
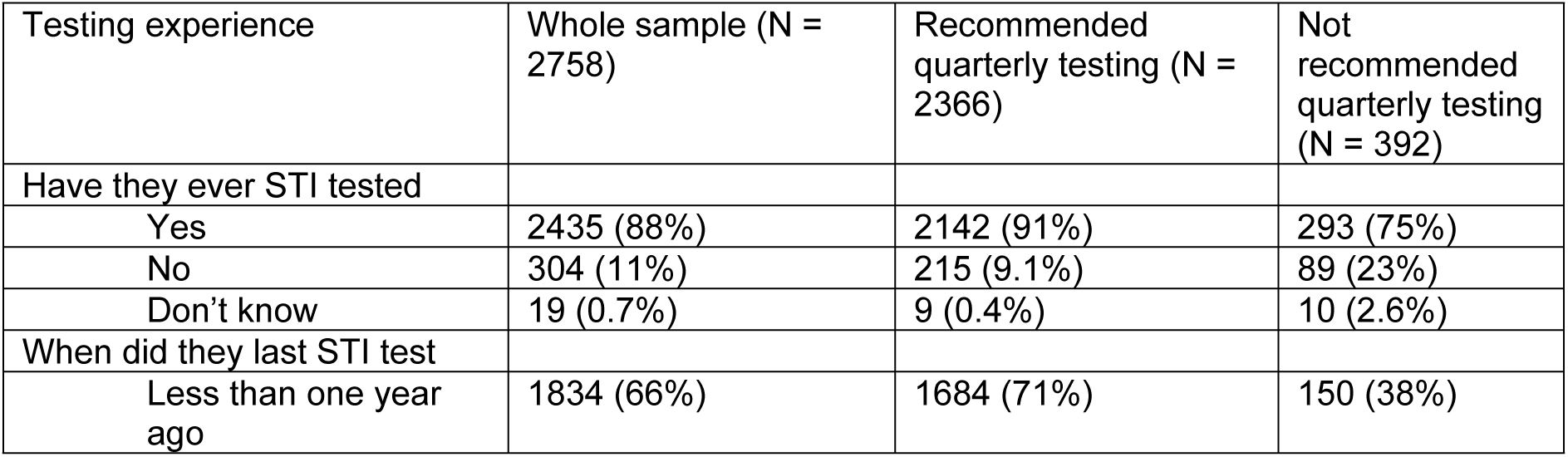
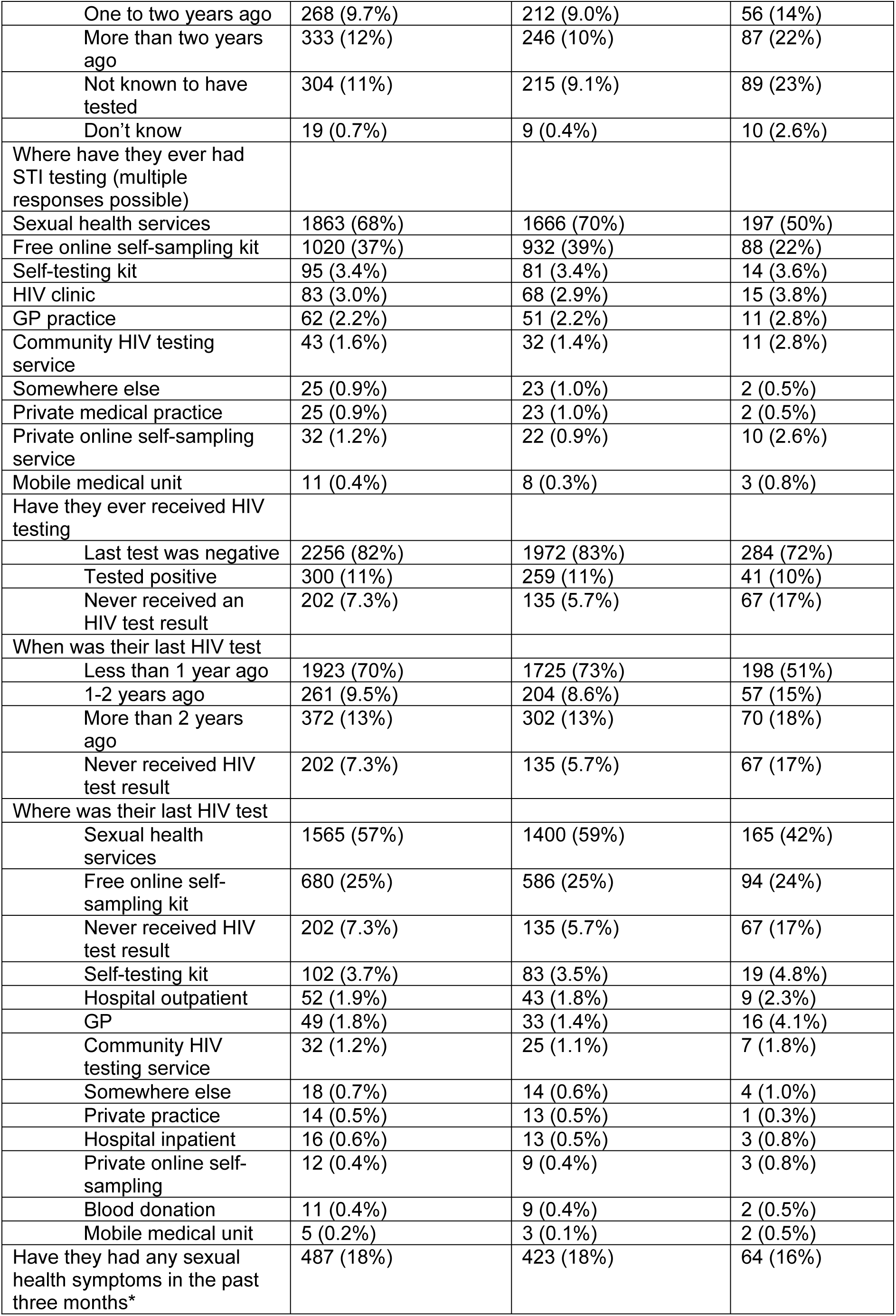
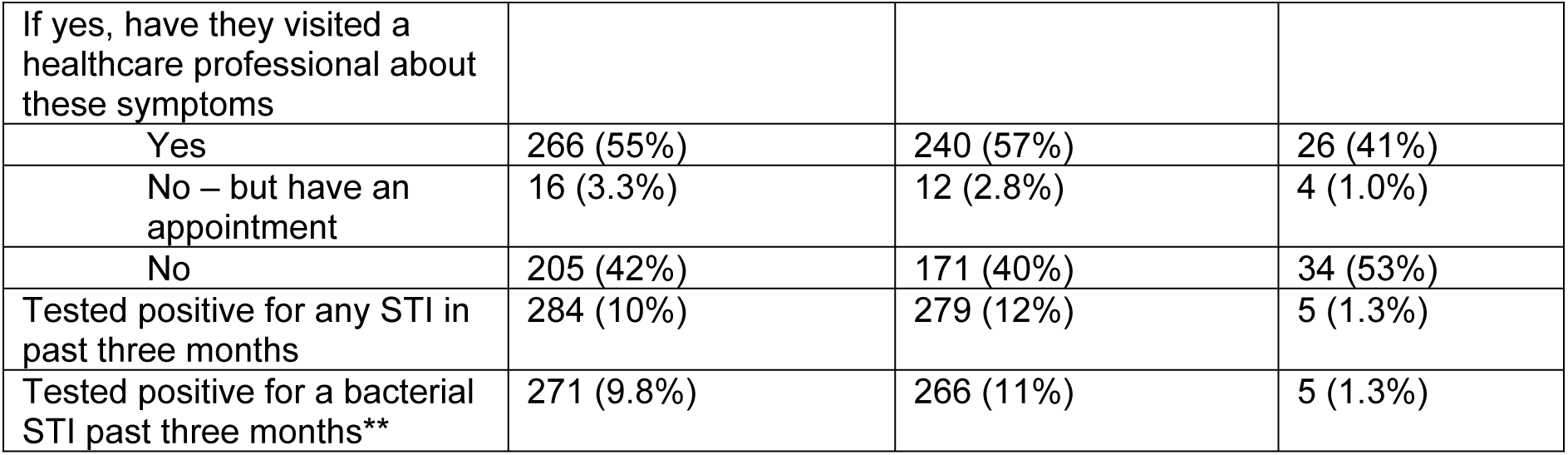
STI and HIV testing experiences reported by the whole sample and those recommended quarterly testing, RiiSH survey, 2024. Sexually transmitted infection (STI) and HIV testing experiences reported by participants in the Reducing Inequalities in Sexual Health (RiiSH) survey 2024. *Full list of symptoms is available in the supplementary material (Supplementary Table S1). **Bacterial STIs were defined as gonorrhoea, chlamydia, or syphilis.

| Testing experience | Whole sample (N = 2758) | Recommended quarterly testing (N = 2366) | Not recommended quarterly testing (N = 392) |
| --- | --- | --- | --- |
| Have they ever STI tested |  |  |  |
| Yes | 2435 (88%) | 2142 (91%) | 293 (75%) |
| No | 304 (11%) | 215 (9.1%) | 89 (23%) |
| Don't know | 19 (0.7%) | 9 (0.4%) | 10 (2.6%) |
| When did they last STI test |  |  |  |
| Less than one year ago | 1834 (66%) | 1684 (71%) | 150 (38%) |
| One to two years ago | 268 (9.7%) | 212 (9.0%) | 56 (14%) |
| More than two years ago | 333 (12%) | 246 (10%) | 87 (22%) |
| Not known to have tested | 304 (11%) | 215 (9.1%) | 89 (23%) |
| Don't know | 19 (0.7%) | 9 (0.4%) | 10 (2.6%) |
| Where have they ever had STI testing (multiple responses possible) |  |  |  |
| Sexual health services | 1863 (68%) | 1666 (70%) | 197 (50%) |
| Free online self-sampling kit | 1020 (37%) | 932 (39%) | 88 (22%) |
| Self-testing kit | 95 (3.4%) | 81 (3.4%) | 14 (3.6%) |
| HIV clinic | 83 (3.0%) | 68 (2.9%) | 15 (3.8%) |
| GP practice | 62 (2.2%) | 51 (2.2%) | 11 (2.8%) |
| Community HIV testing service | 43 (1.6%) | 32 (1.4%) | 11 (2.8%) |
| Somewhere else | 25 (0.9%) | 23 (1.0%) | 2 (0.5%) |
| Private medical practice | 25 (0.9%) | 23 (1.0%) | 2 (0.5%) |
| Private online self-sampling service | 32 (1.2%) | 22 (0.9%) | 10 (2.6%) |
| Mobile medical unit | 11 (0.4%) | 8 (0.3%) | 3 (0.8%) |
| Have they ever received HIV testing |  |  |  |
| Last test was negative | 2256 (82%) | 1972 (83%) | 284 (72%) |
| Tested positive | 300 (11%) | 259 (11%) | 41 (10%) |
| Never received an HIV test result | 202 (7.3%) | 135 (5.7%) | 67 (17%) |
| When was their last HIV test |  |  |  |
| Less than 1 year ago | 1923 (70%) | 1725 (73%) | 198 (51%) |
| 1-2 years ago | 261 (9.5%) | 204 (8.6%) | 57 (15%) |
| More than 2 years ago | 372 (13%) | 302 (13%) | 70 (18%) |
| Never received HIV test result | 202 (7.3%) | 135 (5.7%) | 67 (17%) |
| Where was their last HIV test |  |  |  |
| Sexual health services | 1565 (57%) | 1400 (59%) | 165 (42%) |
| Free online self-sampling kit | 680 (25%) | 586 (25%) | 94 (24%) |
| Never received HIV test result | 202 (7.3%) | 135 (5.7%) | 67 (17%) |
| Self-testing kit | 102 (3.7%) | 83 (3.5%) | 19 (4.8%) |
| Hospital outpatient | 52 (1.9%) | 43 (1.8%) | 9 (2.3%) |
| GP | 49 (1.8%) | 33 (1.4%) | 16 (4.1%) |
| Community HIV testing service | 32 (1.2%) | 25 (1.1%) | 7 (1.8%) |
| Somewhere else | 18 (0.7%) | 14 (0.6%) | 4 (1.0%) |
| Private practice | 14 (0.5%) | 13 (0.5%) | 1 (0.3%) |
| Hospital inpatient | 16 (0.6%) | 13 (0.5%) | 3 (0.8%) |
| Private online self-sampling | 12 (0.4%) | 9 (0.4%) | 3 (0.8%) |
| Blood donation | 11 (0.4%) | 9 (0.4%) | 2 (0.5%) |
| Mobile medical unit | 5 (0.2%) | 3 (0.1%) | 2 (0.5%) |
| Have they had any sexual health symptoms in the past three months* | 487 (18%) | 423 (18%) | 64 (16%) |
| If yes, have they visited a healthcare professional about these symptoms |  |  |  |
| Yes | 266 (55%) | 240 (57%) | 26 (41%) |
| No – but have an appointment | 16 (3.3%) | 12 (2.8%) | 4 (1.0%) |
| No | 205 (42%) | 171 (40%) | 34 (53%) |
| Tested positive for any STI in past three months | 284 (10%) | 279 (12%) | 5 (1.3%) |
| Tested positive for a bacterial STI past three months** | 271 (9.8%) | 266 (11%) | 5 (1.3%) |

We estimated that 2366/ 2578 (86%) would be recommended quarterly STI testing based on reporting any new sexual partner (2146), condomless anal sex with a man (1860), more than 10 sexual partners (594), or chemsex (125) (Figure 1). Of these 2366, 58 (2%) participants reported all four indicators, 528 (22%) reported three, 1129 (48%) reported two, and 651 (28%) reported one.

**Figure 1:**
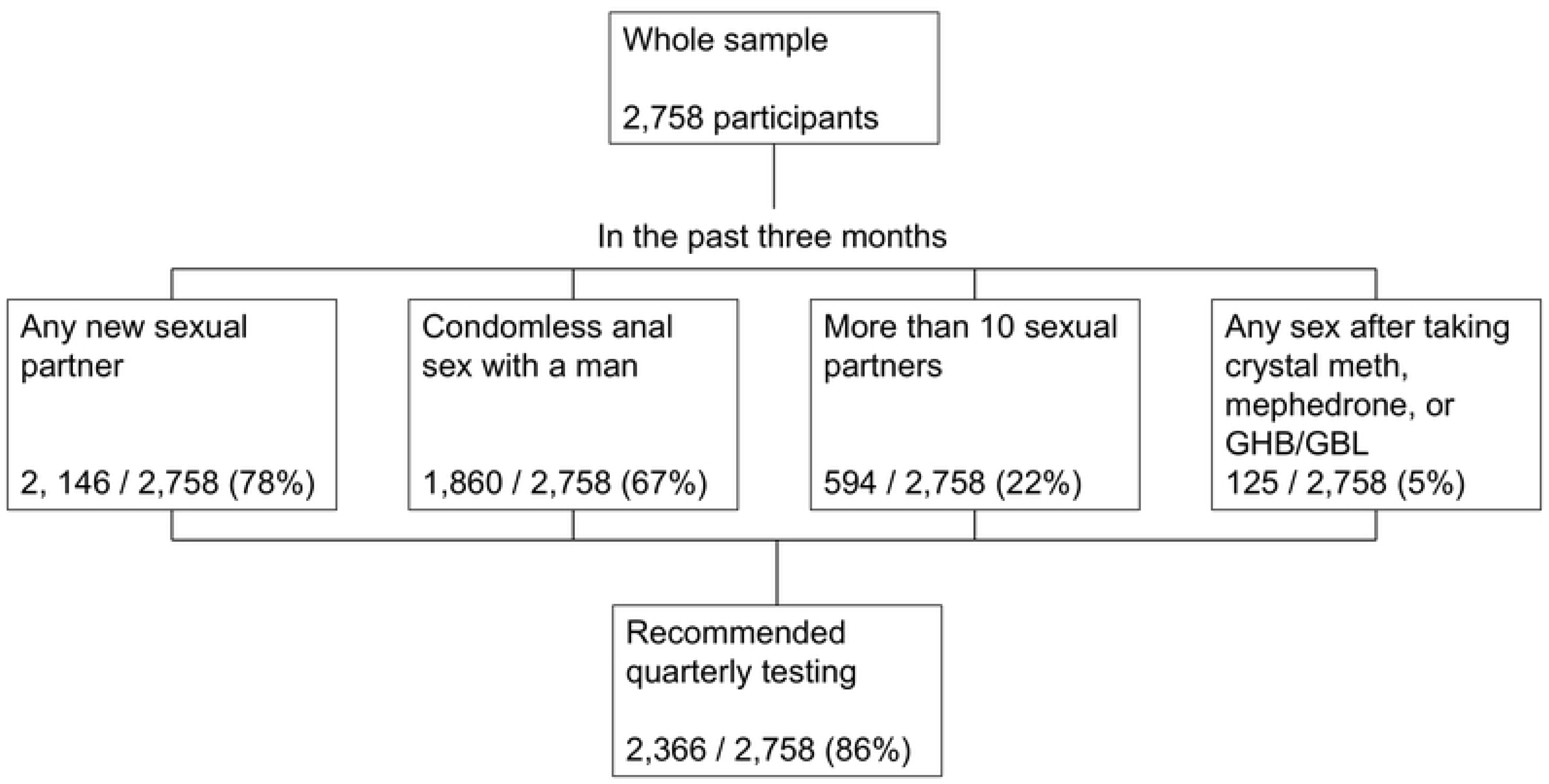
Flowchart describing participants who would be recommended quarterly testing from the RiiSH sample, 2024. Number and proportion of participants in the Reducing Inequalities in Sexual Health (RiiSH) survey 2024 who we estimated would be recommended quarterly testing based on reporting at least one risk factor for STIs over a three-month lookback period (November 2024 looking back to August 2024). The categories are not mutually exclusive and participants could report more than one risk factor.

### STI testing frequency

Of 2366 individuals who would be recommended quarterly testing, where STI testing frequency was available (2342), we found that 562 individuals (24%) reported testing at least four times, 1107 (47%) tested between one and three times, and 673 (28%) did not test in the past year (including 215 (9%) who reported that they had never tested for STIs) (Figure 2). There were 40 (2%) individuals who reported testing at least ten times in the past year.

**Figure 2:**
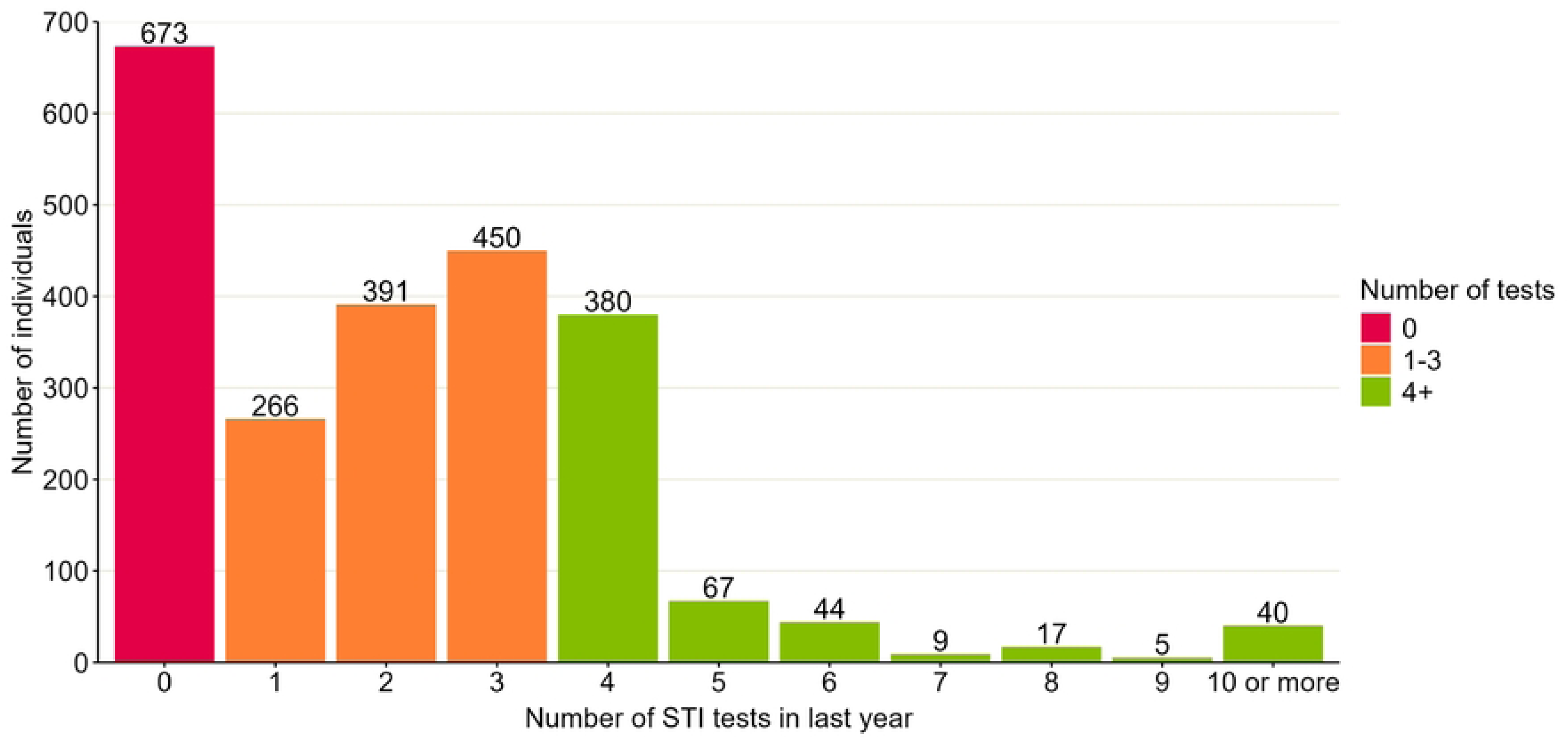
Number of STI tests reported by those RiiSH 2024 survey participants who we estimated would be recommended to test quarterly. Graph includes 2342/2366 participants in the Reducing Inequalities in Sexual Health (RiiSH) survey 2024 who had information available on number of STI tests in the past year. Of 24 without this information, 9 did not know if they had ever STI tested, 11 reported testing in the past year did not remember how many times, and 4 preferred not to say.

### STI and HIV testing experiences

Amongst participants who would be recommended quarterly testing, most reported that they had tested for STIs at least once ever (2142, 91%), and most within the last year (1684, 71%). Attending sexual health services was the most common route that participants had used for STI testing (1666, 70%), followed by using a free online self-sampling kit (932, 39%). Similarly, the majority of participants reported ever having tested for HIV (2231, 96%), with 239 having tested positive (11%). Most participants reported having had an HIV test less than one year ago (1725, 73%). Sexual health services (1400, 59%) and free online self-sampling kits (586, 25%) were how most people had received their last HIV test.

Of those recommended quarterly testing, 423 (18%) reported experiencing any sexual health symptoms in the past three months. Of these, 252/ 423 (60%) reported visiting a healthcare professional about these symptoms or having an appointment booked. Overall, 266 / 2366 (11%) had tested positive for a bacterial STI in the past three months.

### Attendance at sexual health services

As well as questions about STI testing, participants were also asked about attendance at sexual health services, specifically face-to-face appointments. Of 2366 participants recommended quarterly testing, 2083 (88%) had ever visited a sexual health service, 1499 (63%) in the past year, and 1052 (44%) in the past three months (Table 3). The most common reasons for attending a sexual health service were wanting an STI test or general sexual health check-up (57%), to get PrEP (33%), worry about having an STI or HIV without having symptoms (11%), or having symptoms (11%).

**Table 3:**
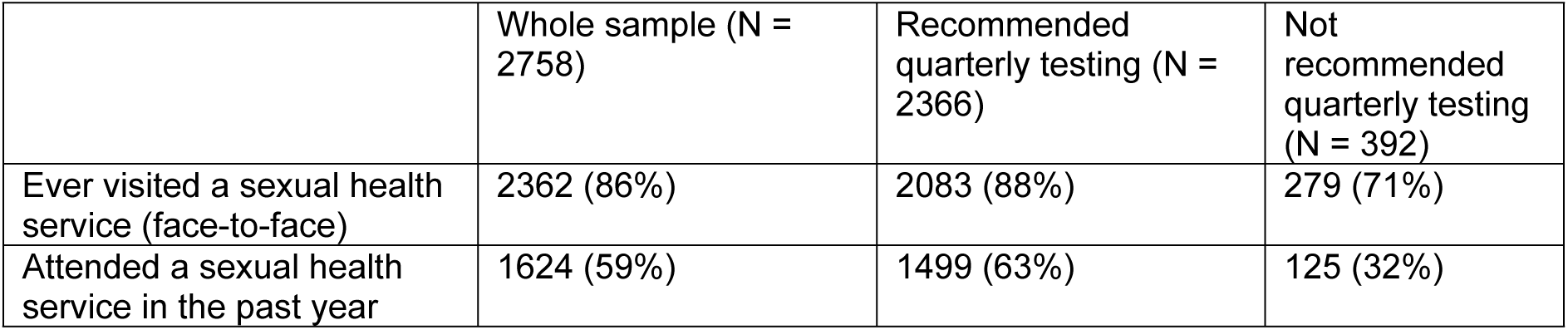
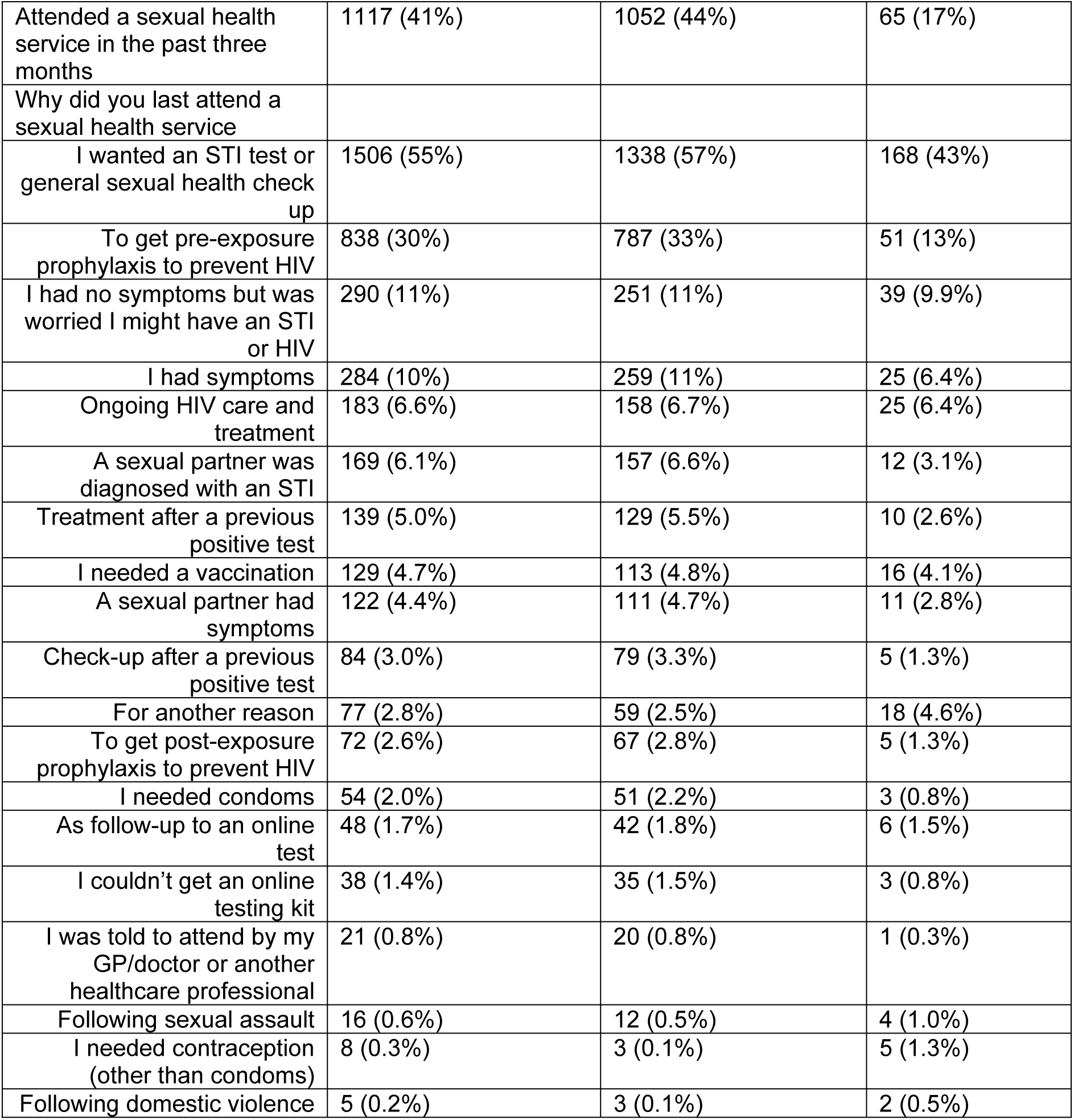
Attendance at sexual health services and reason for attendance, RiiSH survey, 2024. Attendance at sexual health services reported by participants in the Reducing Inequalities in Sexual Health (RiiSH) survey 2024. STI: sexually transmitted infection.

### Correlates of frequent STI testing

The factor most associated with having tested for STIs at least four times in the past year (“frequent testing”) was use of PrEP in the past year (adjusted odds ratio (aOR) 7.66, 95% confidence interval (95% CI) 5.77 - 10.30) (Figure 2, Table 4). Testing STI positive in the last three months (aOR 1.96, 1.45 −2.64) and being aged 16-29 years compared to least 45 years (aOR 1.50, 1.04-2.16), were also associated with frequent testing. Participants who identified as straight or bisexual had lower odds of frequent testing compared to those who identified as gay or homosexual (aOR 0.71, 0.52-0.96).

**Figure 3:**
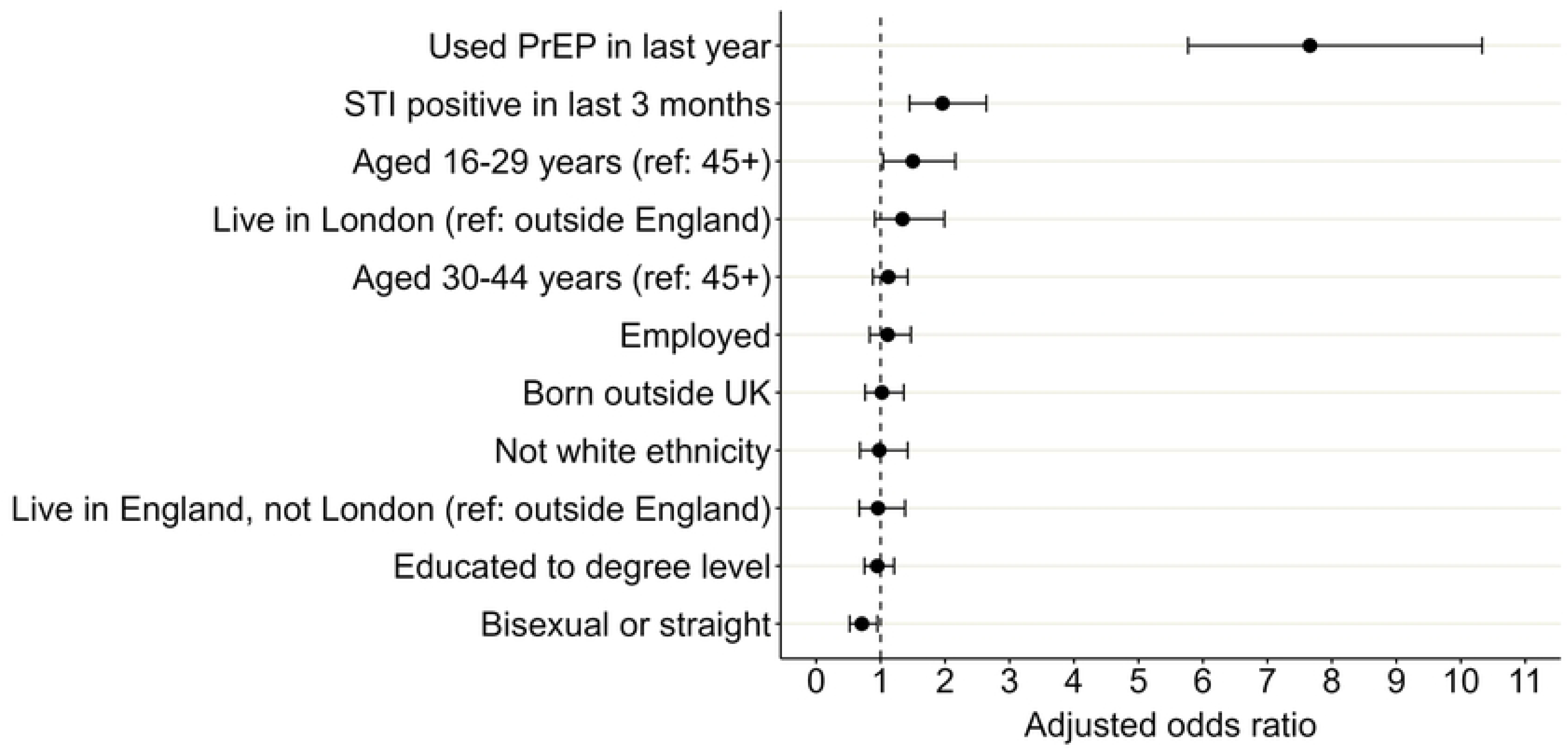
Adjusted odds ratios for factors associated with frequent STI testing (at least four times) compared to those who didn’t test or tested fewer than four times in the past year, RiiSH survey, 2024. Odds ratios showing association of demographic and behavioural factors with frequent testing for sexually transmitted infections (STIs), which was defined as reporting at least four STI tests in the past year, amongst participants in the Reducing Inequalities in Sexual Health (RiiSH) survey 2024. Odds ratios were adjusted for all factors shown in the graph: use of pre-exposure prophylaxis for HIV (PrEP), STI positivity, age, region, employment status, country of birth, ethnicity, level of education, and sexual orientation. Ref = reference group.

**Table 4:** Crude and adjusted odds ratios for factors associated with frequent STI testing (at least four times) compared to those who didn’t test or tested fewer than four times in the past year, RiiSH survey, 2024. Table describes characteristics of individuals who tested for sexually transmitted infections (STIs) frequently (at least four times in the past year) compared to those who did not test or tested less than four times, amongst participants in the Reducing Inequalities in Sexual Health (RiiSH) survey 2024. Table shows crude and adjusted odds ratios (ORs) and confidence intervals (CIs) obtained using multivariable logistic regression, describing the association of demographic and behavioural factors with frequent testing. PrEP: pre-exposure prophylaxis for HIV. Ref: reference group. Adjusted odds ratios were adjusted for all variables shown in the table. Model includes 2,101/ 2,366 observations with complete information. 24 excluded due to missing STI testing frequency because they didn’t know if they had ever tested (9) or how many times they tested (11) or preferred not to say (4). 237 excluded due to missing PrEP use in past year because they had previously tested positive for HIV (227) or did not know if they had ever used PrEP (10). 3 excluded due to not providing information on ethnicity. 8 excluded due to missing information on region of residence within England.

| Characteristic | Frequent<br>(N = 562) |  | Not frequent<br>(N = 1780) |  | Crude |  |  |  | Adjusted |  |  |  |
| --- | --- | --- | --- | --- | --- | --- | --- | --- | --- | --- | --- | --- |
|  | % | n/N | % | n/N | Crude<br>OR | Lower<br>CI | Upper<br>CI | P value | Adjusted<br>OR | Lower<br>CI | Upper<br>CI | P value |
| Used PrEP in last year (ref: no) | 87.5% | 446/ 510 | 44.6% | 714/ 1600 | 8.65 | 6.51 | 11.62 | < 0.001 | 7.66 | 5.77 | 10.30 | <0.001 |
| Tested STI positive in the past three months (ref: no) | 23.3% | 131/ 562 | 8.1% | 144/ 1780 | 3.45 | 2.64 | 4.51 | < 0.001 | 1.96 | 1.45 | 2.64 | <0.001 |
| Sexual orientation (ref: gay/homosexual) | 13.4% | 75/ 562 | 21.1% | 376/ 1780 | 0.58 | 0.43 | 0.76 | < 0.001 | 0.71 | 0.52 | 0.96 | 0.026 |
| Age group (years) |  |  |  |  |  |  |  |  |  |  |  |  |
| 16-29 years | 11.6% | 65/ 562 | 11.5% | 204/ 1780 | 1.15 | 0.84 | 1.56 | 0.375 | 1.50 | 1.04 | 2.16 | 0.029 |
| 30-44 years | 42.0% | 236/ 562 | 35.6% | 633/ 1780 | 1.35 | 1.10 | 1.65 | 0.004 | 1.12 | 0.88 | 1.42 | 0.365 |
| 45+ years (ref) | 46.4% | 261/ 562 | 53.0% | 943/ 1780 | 1.0 | - | - | - | 1.0 | - | - | - |
| Place of residence |  |  |  |  |  |  |  |  |  |  |  |  |
| England – London | 37.6% | 210/ 559 | 25.0% | 444/ 1776 | 1.99 | 1.44 | 2.79 | < 0.001 | 1.34 | 0.91 | 1.99 | 0.147 |
| England – outside London | 52.2% | 292/ 559 | 61.5% | 1092/ 1776 | 1.13 | 0.83 | 1.56 | 0.463 | 0.96 | 0.68 | 1.38 | 0.805 |
| Outside England | 10.2% | 57/ 559 | 13.5% | 240/ 1776 | 1.0 | - | - | - | 1.0 | - | - | - |
| Employment status (ref: not employed) | 81.3% | 457/ 562 | 78.1% | 1390/ 1780 | 1.22 | 0.96 | 1.57 | 0.102 | 1.11 | 0.83 | 1.47 | 0.492 |
| Educated to degree level (ref: other) | 65.7% | 369/ 562 | 59.7% | 1063/ 1780 | 1.29 | 1.05 | 1.58 | 0.012 | 0.95 | 0.75 | 1.21 | 0.668 |
| Born outside UK (ref: born in UK) | 26.7% | 150/ 562 | 20.0% | 356/ 1780 | 1.46 | 1.16 | 1.82 | 0.001 | 1.02 | 0.76 | 1.36 | 0.891 |
| Ethnicity (ref: white) | 11.9% | 67/ 561 | 10.9% | 184/ 1778 | 1.11 | 0.81 | 1.50 | 0.499 | 0.98 | 0.68 | 1.42 | 0.929 |

### Sensitivity analysis

When we compared those who tested at least four times in the past year with those who had tested at least once but fewer than four times, similar risk factors were identified as in the main analysis (Supplementary Table S2). Use of PrEP (aOR 3.99, 2.96-5.45) and having tested positive for an STI (aOR 1.51, 1.13-2.03) were still the factors most strongly associated with frequent testing, but with a lower magnitude of effect than when our reference group included those who hadn’t tested at all in the past year. Younger age (16-29 years compared to over 45 years of age) was also associated with more frequent testing (aOR 1.56, 1.06-2.28), whilst identifying as straight or bisexual was associated with lower odds of frequent testing compared to those who identified as gay or homosexual (aOR 0.72, 0.52-0.98).

## Discussion

As different countries reevaluate their asymptomatic STI testing frequency recommendations, it is important to understand how often individuals are currently testing. Our analyses of data from a large, national community survey of GBMSM suggest that whilst the majority would be recommended to test for STIs every three months, only a quarter met this recommendation. Just under half (47%) reported that they had tested between one and three times, and the remainder (28%) had not tested for STIs in the past year. A small proportion of those eligible for quarterly testing reported that they had never tested for STIs (9%). STI testing had usually taken place at sexual health services or with free self-sampling kits ordered online. Just under half of participants had attended sexual health services for a face-to-face appointment in the past three months. Factors most strongly associated with frequent STI testing were use of PrEP in the last year, STI diagnosis in the past three months, identifying as gay or homosexual rather than straight or bisexual, and younger age.

Our study indicates that not all GBMSM at the highest risk of STIs are testing as often as recommended in national guidelines. Our findings align with previous research in the UK to indicate that testing levels may be lower than expected, with a study in 2019 estimating that GBMSM tested an average of 2.1 times per year [31]. International research has also suggested that testing frequency may be lower than recommended. In Australia, where the guidelines at that time recommended quarterly testing for sexually active GBMSM not in monogamous relationships, a survey of GBMSM in 2021 found that only 31% had tested for STIs at least three times in the past year [32]. Additionally, whilst most GBMSM may be testing at less than the recommended frequency but at least once per year, some are testing less than annually or not at all. A study in the US in 2017 estimated that 34% of GBMSM surveyed at community venues had not tested for STIs in the past year [33]. Our findings are particularly relevant in the current context of uncertainty regarding the benefits and harms of frequent STI testing [15, 21, 34]. Any move to change testing guidance, in the UK or internationally, should consider that the frequency of STI testing amongst GBMSM is likely to be considerably lower than is currently recommended.

We found that PrEP use was strongly associated with frequent STI testing. This is not surprising given that in the UK, PrEP is often provided as a three-month prescription, and requires STI testing [9]. People who choose to use PrEP are self-identifying as having a higher risk of exposure to HIV (and thus to STIs), so it is beneficial that through integration of STI testing with PrEP prescription, those at higher risk are tested more frequently. Similar findings were shown in an Australian survey in 2021, where the proportion of GBMSM who had received STI testing in the past year was much higher amongst those taking PrEP (59%) than those not taking PrEP (8%) [32]. It is also understandable that a recent STI diagnosis would be associated with frequent testing, as at least one test would be required for diagnosis, and possibly further tests of cure. UK guidance goes further to recommend that anyone with a bacterial STI diagnosis in the past year undertakes three-monthly testing [7]. Our findings suggest that these people with greater risk of STIs based on a prior diagnosis are more likely to meet these testing recommendations.

Even though our analysis was restricted to those at risk of STI infection, GBMSM aged over 45 years appeared to be less likely to undergo frequent STI testing. Limited current evidence is available on the relationship between STI testing frequency and age amongst GBMSM. However, studies on the frequency of HIV testing have suggested that older age groups may be less likely to test for HIV, which may also be reflected in STI testing frequency. An English study published in 2014 showed that the odds of not testing for HIV in the past year were greater in GBMSM in their 40s, 50s, or 60s compared to those in their 20s [35]. An American survey published in 2015-2016 also found that older men were less likely to receive comprehensive HIV/STI counselling and testing at their last visit to sexual health services [36]. As well as age, sexual orientation also appears to be associated with STI testing frequency, with straight or bisexual-identifying GBMSM less likely to test frequently. Another research study in the UK reported that MSM identifying as heterosexual were 30% less likely than those identifying as gay or homosexual to have tested for STIs in the past year [37].

Our study utilised a detailed, community-based survey which successfully reached a large number of GBMSM in the UK, many of whom we estimated to have a higher risk of STIs. Our findings give an indication of current STI testing patterns and correlates in this population, providing evidence to feed into decisions on STI testing guidance for GBMSM in the UK. Our study was limited in that as a cross-sectional survey, it could not be used to explore temporality of behaviours in relation to STI testing. We also used proxy definitions for three-monthly testing (describing number of tests in a year rather than timing of tests), and for whether an individual would be recommended quarterly testing (some of whom may not actually be at higher risk, for example, those reporting condomless anal sex within a monogamous relationship). All the data in the survey was self-reported by participants so could be affected by social desirability bias with participants choosing to give desirable answers, which may lead to over-reporting of frequent testing. Due to small numbers, we also binarised some variables (for example, ethnicity and country of birth) and could not explore further the differences between these groups. Finally, RiiSH is not fully representative of the wider GBMSM population, as participants may have a higher risk of STIs due to recruitment through dating applications.

Overall, we found that only 1 in 4 GBMSM with an estimated greater risk of STIs tested as often as recommended. PrEP use, a recent STI diagnosis, and younger age were associated with testing more frequently, whilst straight or bisexual identification was associated with less frequent testing. Any revision to STI testing guidance should take into account that actual testing frequency in GBMSM is likely to be substantially lower than current public health recommendations, and to vary by demographic and behavioural factors.

## Acknowledgements

The authors thank all the participants involved in this study. We acknowledge members of The National Institute for Health and Care Research Health Protection Research Unit (NIHR HPRU) in Blood Borne and Sexually Transmitted Infections (BBSTI) Steering Committee: Professor Caroline Sabin (HPRU Director), Dr John Saunders (PHE Lead), Professor Catherine Mercer, Professor Gwenda Hughes, Professor Greta Rait, Dr Ruth Simmons, Professor William Rosenberg, Dr Tamyo Mbisa, Professor Rosalind Raine, Dr Sema Mandal, Dr Rosamund Yu, Dr Samreen Ijaz, Dr Fabiana Lorencatto, Dr Rachel Hunter, Dr Kirsty Foster and Dr Mamoona Tahir. The authors would like to thank Takudzwa Mukiwa from Terrence Higgins Trust for their help with participant recruitment.

## Data availability statement

The data that support the findings of this study have been assessed by the UK Health Security Agency’s Office for Data Acquisition and Release as having sensitive personal information and are therefore not publicly available to protect participant privacy. However, some aggregate data may be available upon reasonable request from the UKHSA. Requests can be directed to (UKHSA REGG Ref 524).

## Competing interests

The authors have no competing interests to declare.

## Funding

DR, CHM, JS, and HM received funding support as part of The National Institute for Health and Care Research Health Protection Research Unit (NIHR HPRU) in Blood Borne and Sexually Transmitted Infections (BBSTI) at University College London in partnership with the UK Health Security Agency (https://bbsti.hpru.nihr.ac.uk). The funders had no role in study design, data collection and analysis, decision to publish, or preparation of the manuscript. All other authors received no specific funding for this work.

## Authorship

Study design, data collection, and data management were carried out by DO, DR, GB, CHM, JS and HM. DO and HM conceived secondary analysis design with review and contributions from GB, AKH, LF, and DM. LF conducted analyses and wrote the first manuscript draft with contributions from all authors in successive drafts. All authors reviewed and approved the final manuscript.

## Ethical approval

Ethical approval for this study was granted by the UKHSA Research and Ethics Governance Group (REGG; ref: R&D 524) and all methods were performed in accordance with guidelines and regulations set by that group.

## Supplementary Material captions

**S1 Figure title:** Number of male sexual partners in the previous three months reported by participants in the RiiSH survey, 2024

**S1 Figure legend:** Graph showing the number of sexual partners in the previous three months reported by participants in the Reducing Inequalities in Sexual Health (RiiSH) survey 2024

**S1 Table title:** Sexual health symptoms listed in RiiSH 2024 questionnaire

**S1 Table legend:** Sexual health symptoms asked about in the Reducing Inequalities in Sexual Health (RiiSH) survey 2024. Participants were asked different questions about sexual health symptoms depending on the sex they were assigned at birth, which could be AMAB (assigned male at birth), AFAB (assigned female at birth), AIAB (assigned intersex at birth), or PNS (prefer not to say).

**S2 Table title:** Crude and adjusted odds ratios for factors associated with frequent STI testing (at least four times) compared to less frequent testing (one, two, or three times) in the past year, RiiSH survey, 2024

**S2 Table legend:** Table describes characteristics of individuals who tested for sexually transmitted infections (STIs) frequently (at least four times in the past year) compared to those who tested one, two, or three times, amongst participants in the Reducing Inequalities in Sexual Health (RiiSH) survey 2024. Table shows crude and adjusted odds ratios (ORs) and confidence intervals (CIs) obtained using multivariable logistic regression, describing the association of demographic and behavioural factors with frequent testing. PrEP: pre-exposure prophylaxis for HIV. Ref: reference group. Adjusted odds ratios were adjusted for all variables shown in the table. Model includes 1,669/ 1,693 observations which had complete information, where we had excluded 673 participants who had not tested in the past year. 24 were excluded due to missing STI testing frequency because they didn’t know if they had ever tested (9) or how many times they tested (11) or preferred not to say (4). 177 excluded due to missing PrEP use in past year because they had previously tested positive for HIV (175) or did not know if they had ever used PrEP (2). 3 excluded due to not providing information on ethnicity. 4 excluded due to missing information on region of residence within England.

## Notes

### Competing Interest Statement

The authors have declared no competing interest.

### Author Declarations

Ethical approval for this study was granted by the UKHSA Research and Ethics Governance Group (REGG ref: R&D 524) and all methods were performed in accordance with guidelines and regulations set by that group.

## References

1. Mohammed H, Blomquist P, Ogaz D, Duffell S, Furegato M, Checchi M, et al. 100 years of STIs in the UK: a review of national surveillance data. Sex Transm Infect. 2018;94(8):553–8. Epub 20180413. doi: 10.1136/sextrans-2017-053273. PubMed PMID: 29654061.

2. UK Health Security Agency. Spotlight on sexually transmitted infections in London: 2022 data 2024 [11/10/2024]. Available from: https://www.gov.uk/government/publications/sexually-transmitted-infections-london-data/spotlight-on-sexually-transmitted-infections-in-london-2022-data.

3. Stewart J, Baeten JM. HIV pre-exposure prophylaxis and sexually transmitted infections: intersection and opportunity. Nat Rev Urol. 2022;19(1):7–15. Epub 20211025. doi: 10.1038/s41585-021-00527-4. PubMed PMID: 34697493; PubMed Central PMCID: PMCPMC9249100.

4. Gökengin D, Wilson-Davies E, Nazlı Zeka A, Palfreeman A, Begovac J, Dedes N, et al. 2021 European guideline on HIV testing in genito-urinary medicine settings. J Eur Acad Dermatol Venereol. 2021;35(5):1043–57. Epub 20210305. doi: 10.1111/jdv.17139. PubMed PMID: 33666276.

5. Australasian Society for HIV VH, and Sexual Health Medicine (ASHM),. Australian STI management guidelines for use in primary care: Men who have sex with men 2024 [05/06/2025]. Available from: https://sti.guidelines.org.au/populations-and-situations/men-who-have-sex-with-men/.

6. Public Health Agency of Canada. Canadian Guidelines on Sexually Transmitted Infections 2025 [05/06/2025]. Available from: https://www.canada.ca/en/public-health/services/infectious-diseases/sexual-health-sexually-transmitted-infections/canadian-guidelines.html.

7. British Association of Sexual Health and HIV. BASHH Summary Guidance on Testing for Sexually Transmitted Infections, 2023 2023 [11/10/2024]. Available from: https://www.bashh.org/_userfiles/pages/files/resources/bashh_summary_guidance_on_stis_testing_2023.pdf.

8. Clutterbuck D, Asboe D, Barber T, Emerson C, Field N, Gibson S, et al. 2016 United Kingdom national guideline on the sexual health care of men who have sex with men. Int J STD AIDS. 2018:956462417746897. Epub 20180101. doi: 10.1177/0956462417746897. PubMed PMID: 29334885.

9. British HIV Association BAoSHaH. BHIVA/BASHH guidelines on the use of HIV pre-exposure prophylaxis (PrEP), 2018 2018 [28/10/2024]. Available from: https://www.bhiva.org/file/5b729cd592060/2018-PrEP-Guidelines.pdf.

10. Charin G, Symonds Y, Scholfield C, Graham C, Armstrong H. Three-site screening for STIs in men who have sex with men using online self-testing in an English sexual health service. Sexually Transmitted Infections. 2023;99(3):195. doi: 10.1136/sextrans-2021-055243.

11. Danby CS, Cosentino LA, Rabe LK, Priest CL, Damare KC, Macio IS, et al. Patterns of Extragenital Chlamydia and Gonorrhea in Women and Men Who Have Sex With Men Reporting a History of Receptive Anal Intercourse. Sex Transm Dis. 2016;43(2):105–9. doi: 10.1097/olq.0000000000000384. PubMed PMID: 26766527; PubMed Central PMCID: PMCPMC4955797.

12. Mogaka FO, Stewart J, Omollo V, Bukusi E. Challenges and Solutions to STI Control in the Era of HIV and STI Prophylaxis. Curr HIV/AIDS Rep. 2023;20(5):312–9. Epub 20230926. doi: 10.1007/s11904-023-00666-w. PubMed PMID: 37751130; PubMed Central PMCID: PMCPMC10805125.

13. DiNenno EA, Prejean J, Irwin K, Delaney KP, Bowles K, Martin T, et al. Recommendations for HIV Screening of Gay, Bisexual, and Other Men Who Have Sex with Men - United States, 2017. MMWR Morb Mortal Wkly Rep. 2017;66(31):830–2. Epub 20170811. doi: 10.15585/mmwr.mm6631a3. PubMed PMID: 28796758; PubMed Central PMCID: PMCPMC5687782.

14. Rana J, Burchell AN, Wang S, Logie CH, Lisk R, Gesink D. Community perspectives on ideal bacterial STI testing services for gay, bisexual, and other men who have sex with men in Toronto, Canada: a qualitative study. BMC Health Serv Res. 2022;22(1):1194. Epub 20220923. doi: 10.1186/s12913-022-08529-7. PubMed PMID: 36138450; PubMed Central PMCID: PMCPMC9502589.

15. Chow EPF, Fairley CK, Kong FYS. STI pathogens in the oropharynx: update on screening and treatment. Current Opinion in Infectious Diseases. 2024;37(1):35–45. doi: 10.1097/qco.0000000000000997. PubMed PMID: 00001432-202402000-00006.

16. Kenyon C, Herrmann B, Hughes G, de Vries HJC. Management of asymptomatic sexually transmitted infections in Europe: towards a differentiated, evidence-based approach. The Lancet Regional Health – Europe. 2023;34. doi: 10.1016/j.lanepe.2023.100743.

17. van Bergen J, Hoenderboom BM, David S, Deug F, Heijne JCM, van Aar F, et al. Where to go to in chlamydia control? From infection control towards infectious disease control. Sex Transm Infect. 2021;97(7):501–6. Epub 20210527. doi: 10.1136/sextrans-2021-054992. PubMed PMID: 34045364; PubMed Central PMCID: PMCPMC8543211.

18. Tsoumanis A, Hens N, Kenyon CR. Is Screening for Chlamydia and Gonorrhea in Men Who Have Sex With Men Associated With Reduction of the Prevalence of these Infections? A Systematic Review of Observational Studies. Sex Transm Dis. 2018;45(9):615–22. doi: 10.1097/olq.0000000000000824. PubMed PMID: 29485537.

19. Chow EP, Camilleri S, Ward C, Huffam S, Chen MY, Bradshaw CS, et al. Duration of gonorrhoea and chlamydia infection at the pharynx and rectum among men who have sex with men: a systematic review. Sex Health. 2016;13(3):199–204. doi: 10.1071/sh15175. PubMed PMID: 26886136.

20. Barbee LA, Soge OO, Khosropour CM, Haglund M, Yeung W, Hughes J, et al. The Duration of Pharyngeal Gonorrhea: A Natural History Study. Clin Infect Dis. 2021;73(4):575–82. doi: 10.1093/cid/ciab071. PubMed PMID: 33513222; PubMed Central PMCID: PMCPMC8366826.

21. Williams E, Williamson DA, Hocking JS. Frequent screening for asymptomatic chlamydia and gonorrhoea infections in men who have sex with men: time to re-evaluate? Lancet Infect Dis. 2023;23(12):e558–e66. Epub 20230726. doi: 10.1016/s1473-3099(23)00356-0. PubMed PMID: 37516129.

22. Wayal S, Reid D, Weatherburn P, Blomquist P, Fabiane S, Hughes G, et al. Association between knowledge, risk behaviours, and testing for sexually transmitted infections among men who have sex with men: findings from a large online survey in the United Kingdom. HIV Med. 2019;20(8):523–33. Epub 20190524. doi: 10.1111/hiv.12753. PubMed PMID: 31124278; PubMed Central PMCID: PMCPMC6771985.

23. Chandra C, Weiss KM, Kelley CF, Marcus JL, Jenness SM. Gaps in Sexually Transmitted Infection Screening Among Men who Have Sex with Men in Pre-exposure Prophylaxis (PrEP) Care in the United States. Clin Infect Dis. 2021;73(7):e2261–e9. doi: 10.1093/cid/ciaa1033. PubMed PMID: 32702116; PubMed Central PMCID: PMCPMC8492145.

24. Howarth AR, Saunders J, Reid D, Kelly I, Wayal S, Weatherburn P, et al. ’Stay at home …’: exploring the impact of the COVID-19 public health response on sexual behaviour and health service use among men who have sex with men: findings from a large online survey in the UK. Sex Transm Infect. 2022;98(5):346–52. Epub 20210920. doi: 10.1136/sextrans-2021-055039. PubMed PMID: 34544888; PubMed Central PMCID: PMCPMC8457994.

25. Brown JR, Reid D, Howarth AR, Mohammed H, Saunders J, Pulford CV, et al. Changes in STI and HIV testing and testing need among men who have sex with men during the UK’s COVID-19 pandemic response. Sex Transm Infect. 2022;99(4):226–38. Epub 20220721. doi: 10.1136/sextrans-2022-055429. PubMed PMID: 35863887; PubMed Central PMCID: PMCPMC10313956.

26. Ogaz D, Allen H, Reid D, Brown JRG, Howarth AR, Pulford CV, et al. COVID-19 infection and vaccination uptake in men and gender-diverse people who have sex with men in the UK: analyses of a large, online community cross-sectional survey (RiiSH-COVID) undertaken November-December 2021. BMC Public Health. 2023;23(1):829. Epub 20230505. doi: 10.1186/s12889-023-15779-5. PubMed PMID: 37147609; PubMed Central PMCID: PMCPMC10161154.

27. Baldry G, Phillips D, Wilkie R, Checchi M, Folkard K, Simmons R, et al. Factors associated with human papillomavirus, hepatitis A, hepatitis B and mpox vaccination uptake among gay, bisexual and other men who have sex with men in the UK-findings from the large community-based RiiSH-Mpox survey. Int J STD AIDS. 2024;35(12):963–81. Epub 20240820. doi: 10.1177/09564624241273778. PubMed PMID: 39163149.

28. Ogaz D, Edney J, Phillips D, Mullen D, Reid D, Wilkie R, et al. Knowledge, uptake and intention to use antibiotic post-exposure prophylaxis and meningococcal B vaccine (4CMenB) for gonorrhoea among a large, online community sample of gay, bisexual and other men who have sex with men in the UK. medRxiv. 2024:2024.07.08.24310063. doi: 10.1101/2024.07.08.24310063.

29. Ogaz D, Enayat Q, Brown JRG, Phillips D, Wilkie R, Jayes D, et al. Mpox Diagnosis, Behavioral Risk Modification, and Vaccination Uptake among Gay, Bisexual, and Other Men Who Have Sex with Men, United Kingdom, 2022. Emerg Infect Dis. 2024;30(5):916–25. Epub 20240404. doi: 10.3201/eid3005.230676. PubMed PMID: 38573160; PubMed Central PMCID: PMCPMC11060451.

30. Brown JRG, Reid D, Howarth AR, Mohammed H, Saunders J, Pulford CV, et al. Sexual behaviour, STI and HIV testing and testing need among gay, bisexual and other men who have sex with men recruited for online surveys pre/post-COVID-19 restrictions in the UK. Sexually Transmitted Infections. 2023;99(7):467. doi: 10.1136/sextrans-2022-055689.

31. MacGregor L, Speare N, Nicholls J, Harryman L, Horwood J, Kesten JM, et al. Evidence of changing sexual behaviours and clinical attendance patterns, alongside increasing diagnoses of STIs in MSM and TPSM. Sexually Transmitted Infections. 2021;97(7):507. doi: 10.1136/sextrans-2020-054588.

32. Chan C, Holt M, Broady TR, Traeger MW, Mao L, Grulich AE, et al. Trends in Testing and Self-Reported Diagnoses of Sexually Transmitted Infections in Gay and Bisexual Men in Australia, 2017 to 2021: Analysis of National Behavioral Surveillance Surveys. Sexually Transmitted Diseases. 2023;50(12):789–95. doi: 10.1097/olq.0000000000001870. PubMed PMID: 00007435-202312000-00003.

33. Johnson Jones ML, Chapin-Bardales J, Bizune D, Papp JR, Phillips C, Kirkcaldy RD, et al. Extragenital Chlamydia and Gonorrhea Among Community Venue-Attending Men Who Have Sex with Men - Five Cities, United States, 2017. MMWR Morb Mortal Wkly Rep. 2019;68(14):321–5. Epub 20190412. doi: 10.15585/mmwr.mm6814a1. PubMed PMID: 30973847; PubMed Central PMCID: PMCPMC6459584.

34. Berners-Lee W, Cabecinha M, Bell J, Phillips D, Witney T, Pulford CV, et al. ’It does fill me with a bit of unease’: a qualitative study of the acceptability, facilitators and barriers to reducing the frequency of screening for asymptomatic sexually transmitted infections among gay, bisexual and other men who have sex with men. Sex Transm Infect. 2025. Epub 20250715. doi: 10.1136/sextrans-2025-056556. PubMed PMID: 40664501.

35. Witzel TC, Melendez-Torres GJ, Hickson F, Weatherburn P. HIV testing history and preferences for future tests among gay men, bisexual men and other MSM in England: results from a cross-sectional study. BMJ Open. 2016;6(9):e011372. Epub 20160914. doi: 10.1136/bmjopen-2016-011372. PubMed PMID: 27630068; PubMed Central PMCID: PMCPMC5030541.

36. Parsons JT, John SA, Whitfield THF, Cienfuegos-Szalay J, Grov C. Human Immunodeficiency Virus/Sexually Transmitted Infection Counseling and Testing Services Received by Gay and Bisexual Men Using Preexposure Prophylaxis at Their Last PrEP Care Visit. Sex Transm Dis. 2018;45(12):798–802. doi: 10.1097/olq.0000000000000880. PubMed PMID: 30422969; PubMed Central PMCID: PMCPMC6247810.

37. Curtis TJ. The sexual behaviour and sexual health of heterosexual-identifying men who have sex with men: understanding an understudied population to inform public health policy and practice. Doctoral thesis (Ph.D): University College London; 2022.

